# Association of Social Deprivation with Wait Time and Referral Attrition in Obstructive Sleep Apnea

**DOI:** 10.64898/2026.08.13.26360397

**Authors:** Geoffrey McKinnon, Willis H. Tsai, Ada Ip-Buting, Nicole Duff, Gabriel E. Fabreau, Kerry McBrien, Oliver David, Maoliosa Donald, Sachin R. Pendharkar

## Abstract

**Key Points:** *Question:* Among adults referred for specialist obstructive sleep apnea care, is area-level social deprivation associated with referral attrition or wait times for care?

*Findings:* In this retrospective cross-sectional study of 3111 patients, higher situational vulnerability was associated with lower odds of receiving a scheduled appointment, whereas higher ethnocultural composition was associated with higher odds. After an appointment was scheduled, social deprivation was not associated with wait time, cancellations, or no-shows.

*Meaning:* Social inequities in specialist OSA care appear to occur before patients reach the clinic, highlighting completion of referral scheduling pathways as a key intervention to improve equitable access.

**Importance:** Socially vulnerable patients have a high burden of obstructive sleep apnea, but the stage of the referral pathway at which access barriers arise is uncertain.

**Objective:** To determine whether area-level social deprivation was associated with appointment scheduling, wait time, cancellations, or no-shows among adults referred for specialist obstructive sleep apnea care.

**Design:** This was a cross-sectional study evaluating patients referred from December 1, 2016 through November 30, 2019. Data were analyzed from January 30, 2026 to May 16, 2026.

**Setting:** Foothills Medical Centre Sleep Centre in Calgary, Canada.

**Participants:** Adults referred to a tertiary academic sleep centre in Calgary, Alberta, Canada. Eligible patients had valid provincial health insurance and either a scheduled clinic appointment or home sleep apnea test data available.

**Exposures:** Quintiles of the four Canadian Index of Multiple Deprivation domains: residential instability, economic dependency, ethnocultural composition, and situational vulnerability.

**Main Outcomes and Measures:** The primary outcome was receipt of a scheduled specialist appointment. Secondary outcomes were time from referral to the first attended appointment and number of appointment cancellations or no-shows.

**Results:** Among 3111 patients (mean [SD] age, 53.7 [14.3] years; 40.7% female), 1766 (56.7%) were scheduled and 1647 (52.9%) attended an appointment. Each quintile increase in situational vulnerability was associated with lower odds of scheduling (adjusted odds ratio [95% confidence interval] 0.86 [0.81-0.92]), whereas each quintile increase in ethnocultural composition was associated with higher odds (adjusted odds ratio [95% confidence interval] 1.32 [1.22-1.43]). Residential instability and economic dependency were not associated with scheduling. No deprivation domain was associated with time to the first attended appointment, cancellations, or no-shows.

**Conclusions and Relevance:** In this cohort, area-level deprivation was associated with whether patients were scheduled for an appointment but not with wait time or missed visits after scheduling. These findings suggest that equity interventions should focus on completion of referral and scheduling processes.

## Introduction

Obstructive sleep apnea (OSA) is a chronic condition characterized by episodic interruptions in breathing due to recurrent upper airway obstruction during sleep. The estimated prevalence of OSA ranges from 3% to 50%,^1,2^ and an estimated 80% of adults with OSA remain undiagnosed.^3^ Untreated OSA is associated with neurocognitive deficits, excessive daytime sleepiness, cardiovascular disease, and early mortality.^4,5,6^ Treatment of OSA is associated with improved clinical outcomes and is cost-effective compared to no treatment.^7–14^

Socially vulnerable patients are known to have a higher prevalence of OSA than the general population,^15^ and prior studies have suggested that OSA is more severe among socially vulnerable individuals,^16–21^ suggesting that structural and social factors may hinder timely engagement with care. Longer wait times for OSA care have also been associated with decreased treatment adherence and poorer outcomes.^19^ Given the higher prevalence and severity of OSA among socially vulnerable populations, addressing barriers to timely diagnosis and treatment is essential to improving adherence, reducing morbidity, and promoting equity in sleep health.

Prior research has examined the relationship between social vulnerability and wait times for specialist care. Socially vulnerable patients may face access barriers that compound existing challenges such as communication failures between clinicians and patients, and inconsistent information about required documentation or eligibility (e.g., burdensome paperwork prior to appointments).^22,23,24^ Furthermore, these barriers may manifest at multiple stages of the referral process - with a substantial proportion of referrals lost to follow-up, never scheduled, or never attended - leading to gaps in diagnosis and treatment.^25,26^ Social vulnerability has also been associated with greater likelihood of no-shows and cancellations in outpatient settings.^27,28^ There are likely several reasons for this, including transportation barriers, childcare constraints, work inflexibility, and housing instability.^29–32^

To our knowledge, the relationship between social vulnerability, wait times, and patient attrition through the referral pathway for OSA care remains uninvestigated. In this study, we investigated whether social vulnerability influences the probability of being scheduled for an appointment among patients referred for specialist OSA care. Our secondary objectives were to explore the relationship between social vulnerability and overall wait times for OSA care, as well as social vulnerability and cancelled or unattended appointments.

## Methods

### Study Design

This retrospective cross-sectional study evaluated the relationship between social vulnerability and referral outcomes at a tertiary sleep clinic. The study protocol was approved by the University of Calgary Conjoint Health Research Ethics Board (REB-ID 21-0433). This study followed the Strengthening the Reporting of Observational Studies in Epidemiology (STROBE) reporting guideline.^33^

### Study Setting

In Calgary, patients with OSA can be managed by primary care providers (PCPs) using community-based home sleep apnea testing (HSAT) followed by initiation of therapy or be referred to a sleep specialist at the Foothills Medical Centre (FMC) Sleep Centre or in the community. The FMC Sleep Centre is the single tertiary academic sleep centre with expertise in complex sleep-disordered breathing in Southern Alberta, with a catchment area of more than 2.5 million people. Patients who are referred to the FMC Sleep Centre typically undergo HSAT if not completed prior to referral, and complete triage questionnaires. Once these steps are completed, a triage clinician accepts the referral or returns it to the PCP for management based on OSA severity or symptoms. Accepted patients are scheduled for an appointment with a sleep specialist physician.

From November 2016 to February 2017, referrals were accepted regardless of severity criteria. In March 2017, due to capacity constraints at the FMC Sleep Centre and the availability of OSA management by community sleep specialists or PCPs, triage criteria were modified to prioritize patients with severe sleep-disordered breathing (HSAT revealing oxygen desaturation index (ODI) ≥ 30/hour or mean nocturnal oxygen saturation (SpO_2_) < 85%), severe symptoms (Epworth Sleepiness Scale score ≥ 16) or any of the following criteria: self-reported motor vehicle collision within one year, hypertension requiring three or more antihypertensive medications, safety-critical occupation, hospital admission within 30 days prior to referral due to unstable cardiopulmonary or cerebrovascular disease, or upcoming major surgery within 6 months of referral. ODI was used to characterize OSA severity because it was available across a larger proportion of the cohort than AHI and is closely associated with AHI.^34^

Patients with accepted referrals were scheduled for an appointment with a respirologist with sleep medicine training or respiratory therapist with sleep respirologist supervision.^35^

### Cohort Definition

We included all adult patients (age ≥ 18 years) referred to the FMC Sleep Centre for evaluation of OSA from December 1, 2016 to November 30, 2019. Patients were included if they had valid public healthcare insurance coverage under the Alberta Health Care Insurance Plan (AHCIP) at the time of referral. We excluded patients with reason for referral besides OSA or home addresses outside of Alberta. Patients who had neither a scheduled appointment nor HSAT date within the electronic medical record were also excluded as no data were available for analysis. We distinguished these patients from those who were scheduled but cancelled or did not attend appointments, as each group reflects barriers at different points in the care pathway.

### Data Collection

Referral, appointment and clinical data were obtained from the FMC Sleep Centre’s electronic medical record. Variables of interest included patient demographics, referral and appointment dates (scheduled and attended) and sleep testing data. Comorbidity information was derived from administrative databases, including the CIHI Discharge Abstract Database, CIHI National Ambulatory Care Reporting System, Canadian census data, Alberta Health practitioner claims, and Alberta Precision Labs database. Comorbidities were ascertained using previously validated algorithms.^36^

The Canadian Index of Multiple Deprivation (CIMD) is a postal code-derived metric that combines four domains of social vulnerability: residential instability, economic dependency, ethno-cultural composition and situational vulnerability. The CIMD offers a comprehensive and multi-faceted measure of SES that accounts for differential vulnerability across domains.^37^ Patient residential postal codes at the time of referral were linked to geographic dissemination areas using Statistics Canada’s Postal Code Conversion File to obtain census neighbourhood data. Patient-level CIMD domain measures and corresponding quintile classifications were assigned based on the dissemination area linked to each patient’s residential postal code, using geographic indices derived from the 2016 Canadian census. Since the CIMD was designed to capture conceptually different domains of social vulnerability, we analyzed each CIMD domain separately rather than as a composite score.

### Statistical Analysis

Descriptive statistics summarized patient baseline characteristics and time to first appointment. The association between CIMD domain quintile and receipt of a scheduled appointment was assessed using mixed-effects logistic regression with dissemination area as a grouping variable. We calculated adjusted estimates using the following comorbidities: asthma, atrial fibrillation, congestive heart failure, chronic kidney disease, chronic pain, chronic pulmonary disease, dementia, depression, diabetes mellitus, hypertension, prior myocardial infarction, peripheral vascular disease, prior stroke, and hypothyroidism. Mixed-effects linear regression models were constructed to assess the relationships between CIMD domain quintile and wait time for attended specialist appointment adjusted for comorbidities. Patients who did not attend any scheduled appointments were censored from this analysis. We also constructed mixed-effects linear regression models to assess the relationship between CIMD domain quintile and each of the number of appointment cancellations and the number of no-shows. Analyses were performed using Stata Version 17 (StataCorp 2021, College Station, Texas, United States). P values < 0.05 were considered statistically significant.

## Results

We included 3111 referred patients, of whom 1766 (56.7%) were scheduled for an initial clinic appointment. Once scheduled, 1504 patients attended the initial appointment, 198 (11.2% of scheduled patients) cancelled and 64 (3.6% of scheduled patients) did not attend. Of the 262 patients with an initial cancellation or no-show, 143 subsequently attended. Overall, 1647 (52.9% of all referrals, 93.2% of scheduled patients) patients attended a clinic appointment and 119 (3.8% of all referrals, 6.7% of scheduled patients) did not (Figure 1). Home sleep apnea testing data were unavailable for 817, which we attributed to missing data entry for community-based testing. Baseline demographic and clinical characteristics are summarized in Table 1. Mean age was 53.7 (SD 14.3) years and 1266 (40.7%) were female. Patients without scheduled appointments were younger and had lower BMI, less severe OSA and lower comorbidity burden.

**Figure 1.**
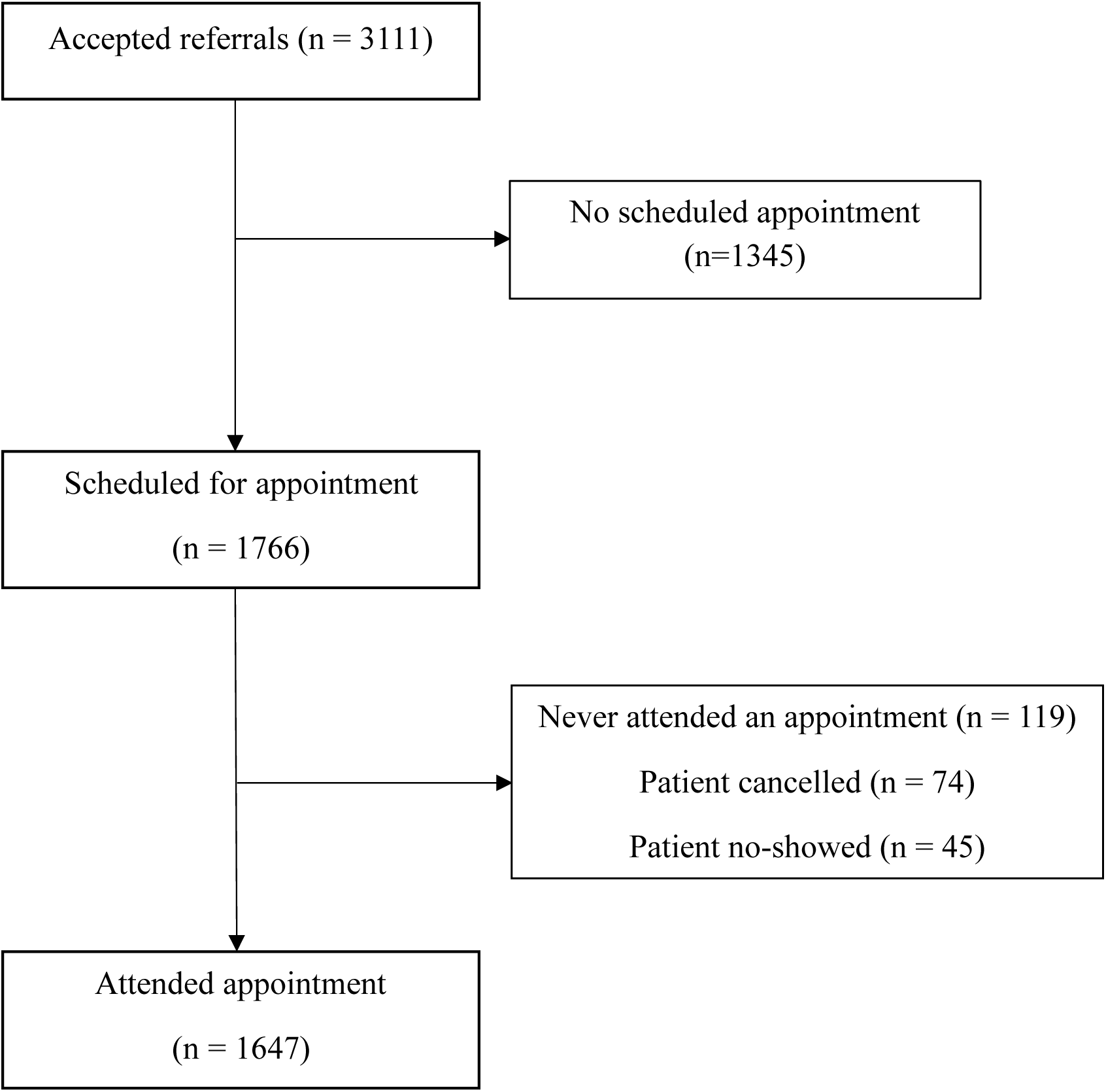
Patient flow from initial referral to attended appointment.

**Table 1.** Baseline Characteristics of Referred Patients.

| Characteristic | All patients<br>(n=3111) | Patients with<br>scheduled<br>appointment<br>(n=1766) | Patients without<br>scheduled<br>appointment<br>(n=1345) |
| --- | --- | --- | --- |
| ***Demographics*** |  |  |  |
| Age, years | 53.7 (14.3) | 57.9 (14.4) | 52.0 (14.5) |
| BMI, kg/m<sup>2</sup>, <sup>a</sup> | 32.2 (7.9) | 36.3 (9.0) | 30.4 (6.9) |
| Male sex, n (%) | 1845 (59.3) | 1056 (59.8) | 789 (58.7) |
| ***OSA Severity<sup>b</sup>*** |  |  |  |
| ODI, events/hr | 18.8 (19.3) | 39.3 (27.3) | 11.1 (9.2) |
| Mean nocturnal<br>SpO<sub>2</sub>, % | 90.7 (3.1) | 87.5 (4.5) | 91.8 (2.0) |
| ***Comorbidities – n (%)<sup>c</sup>*** |  |  |  |
| Asthma | 235 (7.6) | 154 (8.7) | 81 (6.0) |
| Atrial fibrillation | 199 (6.4) | 154 (8.7) | 45 (3.4) |
| Cardiovascular<br>disease | 395 (12.7) | 314 (17.8) | 81 (6.0) |
| Chronic kidney<br>disease | 259 (8.3) | 193 (10.9) | 66 (4.9) |
| Chronic pain | 422 (13.6) | 260 (14.7) | 162 (12.0) |
| Chronic pulmonary<br>disease | 294 (9.5) | 249 (14.1) | 45 (3.4) |
| Depression | 631 (20.3) | 391 (22.1) | 240 (17.8) |
| Diabetes | 683 (22.0) | 517 (29.3) | 166 (12.3) |
| Hypertension | 1265 (40.7) | 883 (50.0) | 382 (28.4) |

Results are presented as mean (standard deviation) unless otherwise specified. Abbreviations: BMI, body mass index; ODI, oxygen desaturation index.
<sup>a</sup> n=2125; <sup>b</sup> n=2294; <sup>c</sup>Comorbidities with at least 5% prevalence in at least one reported column.

Associations between CIMD domains and receipt of a scheduled appointment are presented in Table 2. In unadjusted analyses, each increasing quintile of situational vulnerability was associated with lower odds of having a scheduled appointment (OR 0.81, 95% CI 0.76-0.86, p <0.001), an association that persisted after adjustment (adjusted odds ratio [aOR] 0.86, 95% confidence interval (95% CI) 0.81-0.92, p <0.001). Conversely, each increasing quintile of ethnocultural marginalization was associated with higher odds of having a scheduled appointment in both unadjusted (OR 1.36, 95% CI 1.26-1.47, p <0.001) and adjusted analyses (aOR 1.32, 95% CI 1.22-1.43, p <0.001). Residential instability and economic dependency quintiles were not significantly associated with being scheduled for an appointment.

**Table 2.**
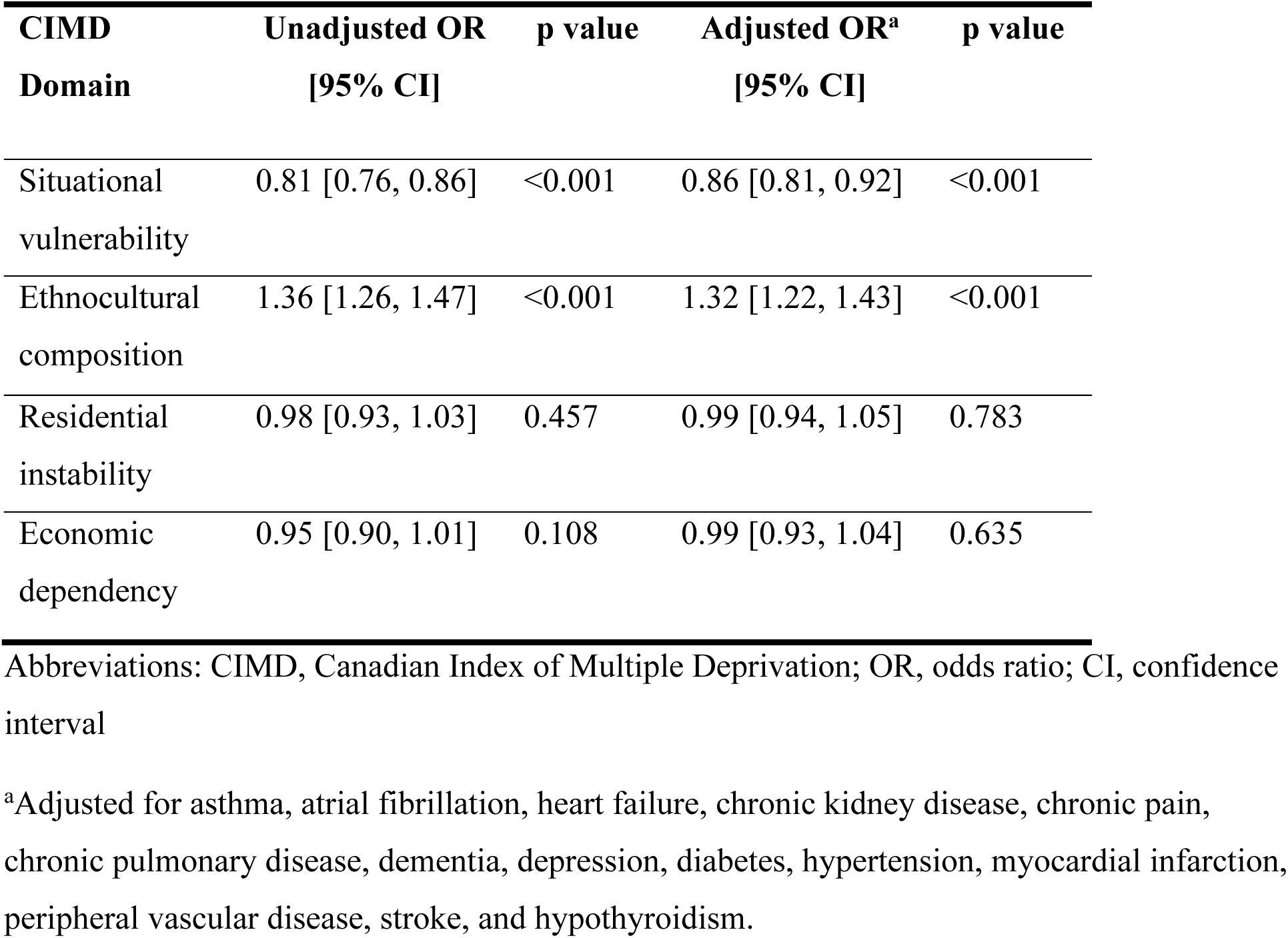
Multivariable logistic regression of the association between CIMD domain quintile and receiving a scheduled appointment (n = 3111).

| **CIMD**<br>**Domain** | **Unadjusted OR**<br>**[95% CI]** | **p value** | **Adjusted OR<sup>a</sup>**<br>**[95% CI]** | **p value** |
| --- | --- | --- | --- | --- |
| Situational<br>vulnerability | 0.81 [0.76, 0.86] | <0.001 | 0.86 [0.81, 0.92] | <0.001 |
| Ethnocultural<br>composition | 1.36 [1.26, 1.47] | <0.001 | 1.32 [1.22, 1.43] | <0.001 |
| Residential<br>instability | 0.98 [0.93, 1.03] | 0.457 | 0.99 [0.94, 1.05] | 0.783 |
| Economic<br>dependency | 0.95 [0.90, 1.01] | 0.108 | 0.99 [0.93, 1.04] | 0.635 |

Abbreviations: CIMD, Canadian Index of Multiple Deprivation; OR, odds ratio; CI, confidence interval
<sup>a</sup>Adjusted for asthma, atrial fibrillation, heart failure, chronic kidney disease, chronic pain, chronic pulmonary disease, dementia, depression, diabetes, hypertension, myocardial infarction, peripheral vascular disease, stroke, and hypothyroidism.

Among patients who were scheduled for an appointment, the mean (standard deviation) time was 86.8 (75.1) days from referral to scheduled appointment and 88.8 (76.2) days from referral to attended appointment. None of the CIMD domains were significantly associated with time from referral to first attended appointment in unadjusted or adjusted analyses (Table 3). Similarly, no CIMD domain was significantly associated with time to first scheduled appointment (data not shown).

**Table 3.** Linear regression of days from referral to first attended appointment by CIMD domain quintile unadjusted and adjusted for comorbidities (n = 1647).

| **CIMD**<br>**Domain** | **Unadjusted**<br>**coefficient**<br>**[95% CI]** | **p value** | **Adjusted**<br>**coefficient**<br>**[95% CI]** | **p value** |
| --- | --- | --- | --- | --- |
| Situational<br>vulnerability | -2.12 [-5.13, 0.89] | 0.167 | -2.14 [-5.19, 0.90] | 0.167 |
| Ethnocultural<br>composition | -0.50 [-3.39, 3.29] | 0.977 | 0.03 [-3.32, 3.37] | 0.987 |
| Residential<br>instability | 0.81 [-1.90, 3.52] | 0.559 | 0.40 [-2.32, 3.13] | 0.773 |
| Economic<br>dependency | 0.21 [-2.70, 3.12] | 0.888 | 0.20 [-2.72, 3.13] | 0.892 |

Abbreviations: CIMD, Canadian Index of Multiple Deprivation; CI, confidence interval
<sup>a</sup>Adjusted for asthma, atrial fibrillation, heart failure, chronic kidney disease, chronic pain, chronic pulmonary disease, dementia, depression, diabetes, hypertension, myocardial infarction, peripheral vascular disease, stroke, and hypothyroidism.

Severe OSA and severe sleepiness were both independently associated with shorter wait times when adjusted for comorbidities. Specifically, patients with an ODI ≥ 30 events/hr were assessed 13.8 days sooner (95% CI −24.60, −3.07) compared to patients with an ODI < 30 events/hr (p=0.012), and patients with an Epworth Sleepiness Scale (ESS) score ≥ 16 were assessed 21.9 days sooner (95% CI −33.27, −10.48) compared to patients with a score < 16 (p <0.001).

The total proportion of appointment cancellations and no-shows was 8.4% of all referrals. In mixed-effects regression models, none of the four CIMD domains was significantly associated with the number of cancelled or unattended clinic appointments.

## Discussion

In this cohort of patients referred for OSA assessment at a tertiary sleep centre in Alberta, Canada, we analyzed the association between dimensions of social vulnerability and access to specialist care at several points in the referral pathway. We found significant patient attrition throughout, with only 56.7% of patients who were originally referred successfully scheduling a specialist appointment. We also found that patients living in areas of higher situational vulnerability were less likely to schedule an appointment. In contrast, ethnocultural composition was associated with a greater tendency to receive a scheduled appointment. None of the CIMD domains were associated with time to first attended appointment, appointment cancellations, or unattended visits. These findings suggest that among patients who are referred for sleep specialist care, disparities in access are concentrated at the point of entry into the specialist system rather than in the delivery of care once patients are engaged.

Social vulnerability has been linked to poor access in other contexts, including in wait times for cancer care, specialty follow-up, initial specialist assessment, and emergency care.^38–41^ Internationally, studies have also demonstrated that social vulnerability is generally associated with longer specialist wait times.^42–44^ Our analysis did not show an association between area-level social vulnerability and wait times for patients with scheduled appointments, which is consistent with other studies that restricted analysis to patients who have already entered a referral pathway for specialty care.^45,46^ However, analyses of wait times among patients who are successfully scheduled may be prone to selection bias, as socially vulnerable patients who encounter the greatest barriers to care may be disproportionately excluded before wait time can be observed. Consistent with this possibility, we observed greater referral attrition in this group, indicating that vulnerable patients encounter obstacles earlier in the pathway that prevent them from ever being scheduled for an appointment. These findings support measurement of referral attrition to evaluate access to care for socially vulnerable populations.

In our study, patients from areas with greater situational vulnerability were less likely to schedule an appointment. Prior research has identified barriers to OSA care that could explain this, including lower health literacy and limited familiarity with medical forms that may hinder completion of sleep questionnaires that are often required for referral acceptance. Additionally, competing priorities such as childcare may make appointment attendance difficult, and lack of reliable transportation may present additional hurdles.^15–18,47,48^ Our results suggest that these structural barriers may contribute to early attrition before clinical contact is established rather than after appointments have been scheduled.

In contrast, we found that patients living in areas of higher ethnocultural composition were more likely to receive a scheduled appointment. This finding was unexpected, as previous research has demonstrated that ethnoculturally marginalized groups face barriers obtaining specialty care for other conditions.^49–51^ One possible explanation is that our multivariable models simultaneously adjusted for the other CIMD domains. As a result, the observed association may reflect ethnoculturally diverse populations who have lower residential instability, economic dependency, or situational vulnerability owing to other factors, such as urban residence or the “healthy immigrant effect,” whereby recent immigrants tend to have better overall health than the non-immigrant population.^52^ It is also possible that unmeasured confounding factors, including differences in healthcare utilization patterns or community-level supports, may mitigate barriers to care. Importantly, our cohort represents patients who had already been referred for specialist assessment and thus may differ from the general population. Finally, these associations may reflect individual characteristics rather than neighbourhood level vulnerability scores.

When the analysis was restricted to patients who were scheduled for an appointment, only higher ODI and ESS scores were associated with shorter wait times, as would be expected based on triage criteria at the FMC Sleep Centre. In contrast, area-level social deprivation did not predict care delays, suggesting that once triage requirements were completed, wait times to access specialist OSA care were not impacted by social vulnerability. Two factors may explain this.

First, referral bias at a tertiary centre may dilute the effects of social deprivation. Referred patients often have more severe disease and greater contact with healthcare, which may outweigh social vulnerability in determining appointment timing within a standardized referral system.

Patients with more severe disease may also engage more consistently with care, even among socially vulnerable groups, and thus be more likely to seek timely care. Referral bias may also be due to the private nature of OSA treatment in Alberta, which may deter pursuit of OSA testing and treatment among those with financial or other barriers to care.^53^ In this regard, our results may reflect systematic exclusion of vulnerable members of the population.

We also observed no association between CIMD domains and either appointment cancellations or no-shows. This contrasts with previous literature in which area-level deprivation was associated with missed healthcare visits.^54^ In sleep medicine specifically, Cheung *et al.* found higher no-show rates among patients with Medicaid or no insurance, which are associated with socioeconomic disadvantage.^55^ Our negative finding may be explained by two factors. First, relatively few patients had a no-show (83 of 1766 scheduled patients [4.7%]), limiting power to detect modest differences by deprivation that are apparent in settings with higher baseline non-attendance. Second, because most referred patients completed HSAT prior to referral or as part of the triage process, our cohort may have been biased to include individuals who are more likely to keep appointments. Finally, greater disease severity and prior engagement with the healthcare system among referred patients may also reduce the influence of social vulnerability on appointment attendance, with patients with more severe disease being more likely to engage with care.

Our study has several strengths. We leveraged a large referral cohort with robustly collected data on referrals, HSAT completion, appointment scheduling, and visit attendance, allowing detailed characterization of attrition across multiple stages of the referral pathway. In addition, we used the Canadian Index of Multiple Deprivation, a well-validated measure of social vulnerability, which enabled assessment of conceptually distinct socioeconomic dimensions with methodological rigor.

However, this study also has important limitations. First, the triage process at the FMC Sleep Centre requires that patients complete HSAT before being scheduled for an appointment, so we assumed that missing data reflected HSAT performed at external sites that were not captured in our database. However, we cannot confirm that HSAT was performed in all patients. Moreover, the extent of missing HSAT data limited our ability to fully account for disease severity in our analysis. Second, changes in triage criteria during the study period resulted in return of referrals for patients with less severe OSA, which may have influenced scheduling more than social vulnerability. While greater social vulnerability might be expected to be associated with more severe disease and therefore a higher likelihood of scheduling, we observed no association between situational vulnerability and OSA severity after adjustment for BMI in a prior analysis using the same cohort.^20^ Thus, we believe that socially vulnerable patients did not have more severe OSA requiring prioritization, and that different disease severity among socially vulnerable patients is unlikely to explain the observed scheduling patterns. Third, although we used mixed-effects regression to account for clustering by dissemination area, area-level analyses remain at risk of ecological fallacy, and neighbourhood-level measures cannot fully substitute for individual-level socioeconomic information. Finally, our cohort does not capture the most vulnerable individuals who never enter the referral pathway because of financial, social, or logistical barriers. Consequently, our finding of no association between social vulnerability and wait times may represent a best-case estimate among patients who successfully entered the referral pathway, as the exclusion of the most vulnerable individuals may have biased associations toward the null. Despite these limitations, the consistency of our findings supports the conclusion that socially vulnerable patient populations are at risk of referral attrition from specialist OSA care.

### Conclusion

In this tertiary care referral cohort, patients with OSA residing in more socially vulnerable neighborhoods were less likely to obtain specialist care. Among patients who were scheduled for an appointment, those with more severe OSA were assessed sooner, and social vulnerability was not associated with delays for care or missed appointments. These findings suggest that triage requirements present a barrier to specialist care for OSA for socially vulnerable patients. Future research should explore interventions to reduce these disparities and improve referral pathway completion among vulnerable populations.

## Article Information

**Author contributions:** Drs. McKinnon, Pendharkar, and Tsai had full access to the data in the study and take responsibility for accuracy of the data and analysis.

*Concept and design:* GM, WHT, ND, SRP

*Acquisition, analysis, or interpretation of data:* GM, WHT, AIB, ND, SRP

*Drafting of manuscript:* GM *Critical revision:* All authors *Statistical analysis: WHT*

*Obtained funding: WHT AIB, GEF, KM, OD, MD, SRP Supervision: SRP*

## Data Availability

All data produced in the present study are available upon reasonable request to the authors

